# Influence of Body Position on the Motor Development of Preterm Infants: A Randomized Clinical Trial

**DOI:** 10.1101/2023.03.01.23285519

**Authors:** Vitória Regina de Morais Cardoso Rodrigues, Rita Cordovil, Marisa Afonso de Andrade Brunherotti, Marisa Afonso Andrade Brunherotti

## Abstract

**Background:** To analyze the influence of body position on the motor development of preterm infants in the first year of life corrected for prematurity.

**Methods:** This controlled, randomized, open trial included 30 preterm infants randomly assigned to one of the following three groups: prone group (n = 9), supine group (n = 10), and control group (n = 11). Intervention: Motor development was assessed at four time points using the Alberta Infant Motor Scale: first outpatient visit and at 4, 8 and 12 months corrected age.

**Results:** In the third assessment at 8 months corrected age, the supine group exhibited better motor development than the other groups (p = 0.02). In the control group, the number of infants with normal development decreased from 11 (100%) in the first assessment to 5 (45.45%) in the last assessment. Most infants of mothers who received guidance on body positioning achieved normal motor development in the first year of life (63.1%). Greater dispersion from normal Alberta Infant Motor Scale scores was observed in infants at 8 and 12 months of age.

**Conclusions:** Guidance on body positioning of preterm infants at home appears to have a positive influence in the first year of life. Child care strategies after hospitalization should be supported to permit full development of the child.

**What’s New:** This study reinforces home care programs with specialized orientation for motor development of preterm newborns. By the end of the first year of life most infants in the experimental groups (supine and prone positions) achieved a normal motor development, which did not happen in the control group.

## Introduction

Preterm infants are a vulnerable group that is an increased risk of physical, neurological and cognitive problems whose severity is inversely proportional to gestational age and the quality of perinatal care.^1,2,3^ Most of the outcomes in this group have psychosocial, emotional and economic impacts on the families, on society, and on the health system. ^4,5^ Within this context, prematurity and low birth weight are biological risk factors for alterations in motor development, especially when the child is born at a gestational age less than 32 weeks and birth weight less than 1,500 g.^6^

For the acquisition of motor developmental milestones, it is important to stimulate newborns in body positions that require greater muscle strength against the action of gravity, such as the prone position.^7^ However, prone positioning after the newborn infant has been discharged home is avoided because it is associated with death from suffocation.^8^

The lack of attention to stimulating different body positions in newborn infants can compromise child development. Studies have shown an association between prone positioning only for short periods and delayed motor development.^9,10^ It is known that Brazilian mothers seldom stimulate prone positioning and offer their lap a lot to their babies, giving them few opportunities to sit on the floor.^11,12^ Thus, studies are needed to better understand this topic within the context of parental habits and practices.^13^

Attention to the development of children in the first years of life is essential since this period is characterized by major modifications and the acquisition of motor skills. The longitudinal follow-up of children using assessment scales allows the early identification of motor disorders and delays and enables early interventions. However, intervention and longitudinal studies of preterm infants in the outpatient setting are still sparse in Brazil.^14^

Therefore, the objective of this study was to analyze the influence of body position on the motor development of preterm infants in the first year of corrected age under home follow-up.

## Method

This was a controlled, randomized, open trial. The study was approved by the Ethics Committee of the University and is registered with the Clinical Trials Registry.

### Participants

Preterm infants (gestational age < 37 weeks) in their first outpatient visit were eligible. Infants with evident neurological and/or orthopedic alterations, malformations, syndromes, confirmed congenital infections, sensory deficits (vision or hearing), chromosome abnormalities, CNS malformations, polymalformative syndromes, congenital heart diseases, severe intraperiventricular hemorrhage, and parenchymal hemorrhagic infarction were excluded.

The minimum sample size was calculated assuming a desired level of confidence of 95% and maximum error of 2.0 for a standard deviation of 5.0 for the percentile score of the Alberta Infant Motor Scale (AIMS). Thus, a minimum number of 25 preterm infants was estimated for this study.

The infants were randomized into the following groups using sealed and opaque envelopes: control group consisting of 11 infants, supine group consisting of 10 infants, and prone group consisting of 10. The infants were evaluated considering the corrected age ^15^ in the first outpatient visit (mean age of 39.27 ± 2.64 weeks), at 4 and 8 months, and when they had completed one year. The assessments were performed by a single evaluator.

### Assessment Instruments

Demographic, clinical and prenatal data of the mothers were collected. In the infants, anthropometric measurements were obtained and motor development was evaluated using the AIMS. The mean time of application of the scale was 20 minutes.

The AIMS measures functional skills and movement quality from birth to 18 months of age.^16^ This scale consists of 58 items divided into four subscales: prone (21 items), supine (9 items), sitting (12 items), and standing (16 items). Each motor skill item is scored as 1 (observed) or 0 (not observed). The total raw score is obtained by summing the scores of each item of the four subscales. The total score and corrected age determine the position of the infant on the norm-referenced percentage curves and motor development is categorized into percentiles: normal/expected (> 25^th^), suspicious (25^th^ to 5^th^), and abnormal (< 5^th^)^.17^

### Procedures and Intervention

In the first assessment after randomization, the parents were asked to implement the instructions on body positioning at home during the waking hours of the infant from the first assessment until the infant began to crawl. The intervention positions (prone and supine) should be applied for 2 hours in the morning and 2 hours in the afternoon in a playful and supervised manner, while at night the position could be chosen freely. The control group received no type of guidance.

In the interval between assessments, the mothers were contacted by telephone or personally in the waiting room of the clinic, providing feedback on the instructions established at the beginning of the study. The mothers also received a booklet to record the days when they followed the instructions of the intervention.

### Statistical Analysis

Statistical analysis was performed using the Statistical Package for the Social Sciences (SPSS) 23.0 for Windows. The maternal characteristics and profile of the preterm infants are reported descriptively using percentage, mean, and standard deviation. Significant differences between the groups were investigated using ANOVAs and Fisher exact tests. Motor gain in the first year of life for each group of the preterm infants was investigated using Friedman’s test and the Wilcoxon post-hoc test. Stepwise multiple linear regressions were performed to identify the best predictors of performance in the AIMS in each assessment. The level of significance was set at p < 0.05.

## Results

During the study period, 45 preterm infants started longitudinal outpatient follow-up and 31 were eligible for the study. One infant was excluded during the intervention because of a diagnosis of Dandy-Walker syndrome. Thus, 30 preterm infants participated in this study.

The maternal characteristics are shown in Table 1. Antenatal corticosteroid use was the only variable that differed significantly between the groups studied, with 90.9% of mothers of the control group using corticosteroids (p = 0.05). There was no significant difference in the other maternal variables between the three groups.

**Table 1.** Distribution of the characteristics of mothers of the infants participating in the study

| Variable | Total<br>group<br>(n = 30)<br>Mean±SD | Control<br>group<br>(n = 11)<br>Mean±SD | Prone<br>group<br>(n = 9)<br>Mean±SD | Supine<br>group<br>(n = 10)<br>Mean±SD | F<br>(2,27) | p |
| --- | --- | --- | --- | --- | --- | --- |
| Maternal age (years) | 27.8±5.9 | 30.2±6.6 | 25.0±4.8 | 27.7±5.1 | 2.13 | 0.14 |
| Prenatal visit | 06.1±2.2 | 06.2±1.4 | 07.0±2.8 | 05.1±2.0 | 1.90 | 0.17 |
|  | n % | n % | n % | n % | FET | p |
| Maternal education |  |  |  |  |  |  |
| Elementary school | 09 (30) | 05 (45.1) | 02 (22.2) | 02 (20.0) |  |  |
| High school | 17 (56.6) | 05 (45.1) | 06 (66.7) | 06 (60.0) | 2.39 | 0.74 |
| Higher education | 04 (13.3) | 01 (9.0) | 01 (11.1) | 02 (20.0) |  |  |
| Type of delivery |  |  |  |  |  |  |
| Cesarean | 24 (80) | 10 (90.9) | 06 (66.7) | 08 (80.0) | 1.81 | 0.38 |
| Normal | 06 (20) | 01 (9.1) | 03 (33.3) | 02 (20.0) |  |  |
| Parity |  |  |  |  |  |  |
| Primiparous | 14 (46.6) | 06 (54.6) | 02 (22.2) | 06 (60.0) | 3.07 | 0.25 |
| Multiparous | 16 (53.3) | 05 (45.4) | 07 (77.8) | 04 (40.0) |  |  |
| Antibiotic use |  |  |  |  |  |  |
| Yes | 12 (40) | 03 (27.3) | 04 (44.4) | 05 (50.0) | 1.29 | 0.64 |
| No | 18 (60) | 08 (72.7) | 05 (55.6) | 05 (50.0) |  |  |
| Antenatal |  |  |  |  |  |  |
| corticosteroid | 19 (63.4) | 10 (90.9)* | 05 (55.6) | 04 (40.0) | 6.22 | 0.05* |
| Yes | 11 (36.6) | 01 (9.1) | 04 (44.4) | 06 (60.0) |  |  |
| No |  |  |  |  |  |  |

Note. FET: Fischer's Exact Test; SD: standard deviation.
\* Significant at $p \leq 0.05$

Table 2 shows the main birth characteristics of the infants studied. No significant difference was observed between the three groups in any of the variables and the groups were therefore considered homogeneous. The mean gestational age of the preterm infants was 30.6±59 weeks and the mean birth weight was 1,367.5±413 kg.

**Table 2.**
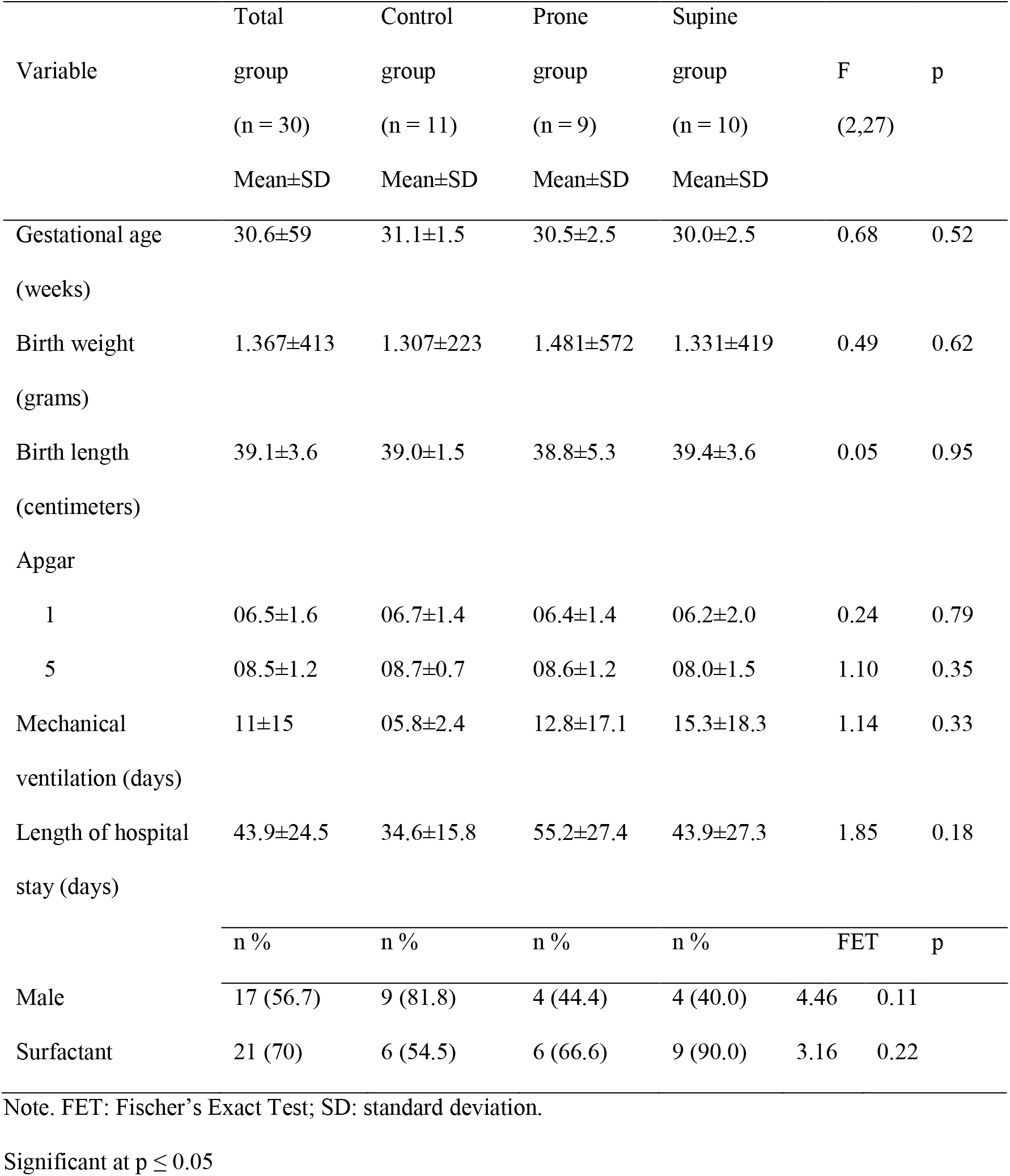
Profile of the 30 preterm infants participating in the study

|  | Total | Control | Prone | Supine |  |  |
| --- | --- | --- | --- | --- | --- | --- |
| Variable | group | group | group | group | F | p |
|  | (n = 30) | (n = 11) | (n = 9) | (n = 10) | (2,27) |  |
|  | Mean±SD | Mean±SD | Mean±SD | Mean±SD |  |  |
| Gestational age | 30.6±59 | 31.1±1.5 | 30.5±2.5 | 30.0±2.5 | 0.68 | 0.52 |
| (weeks) |  |  |  |  |  |  |
| Birth weight | 1.367±413 | 1.307±223 | 1.481±572 | 1.331±419 | 0.49 | 0.62 |
| (grams) |  |  |  |  |  |  |
| Birth length | 39.1±3.6 | 39.0±1.5 | 38.8±5.3 | 39.4±3.6 | 0.05 | 0.95 |
| (centimeters) |  |  |  |  |  |  |
| Apgar |  |  |  |  |  |  |
| 1 | 06.5±1.6 | 06.7±1.4 | 06.4±1.4 | 06.2±2.0 | 0.24 | 0.79 |
| 5 | 08.5±1.2 | 08.7±0.7 | 08.6±1.2 | 08.0±1.5 | 1.10 | 0.35 |
| Mechanical | 11±15 | 05.8±2.4 | 12.8±17.1 | 15.3±18.3 | 1.14 | 0.33 |
| ventilation (days) |  |  |  |  |  |  |
| Length of hospital | 43.9±24.5 | 34.6±15.8 | 55.2±27.4 | 43.9±27.3 | 1.85 | 0.18 |
| stay (days) |  |  |  |  |  |  |
|  | n % | n % | n % | n % | FET | p |
| Male | 17 (56.7) | 9 (81.8) | 4 (44.4) | 4 (40.0) | 4.46 | 0.11 |
| Surfactant | 21 (70) | 6 (54.5) | 6 (66.6) | 9 (90.0) | 3.16 | 0.22 |

Note. FET: Fischer's Exact Test; SD: standard deviation.
Significant at $p \leq 0.05$

Classification of the groups studied by the AIMS in the four assessments (Table 3) showed a significant difference in the third assessment (p = 0.02). In this assessment (8 months of age), the supine group exhibited the best motor development, with 80% of the infants being classified as normal. In the first assessment, all infants were classified as normal. The percentage of infants classified as suspicious and abnormal tended to increase over the phase of motor development Friedman’s test revealed that this increase was only significant in the control group (χ^2^=12.91, p<0.01). The Wilcoxon signed-ranks test indicated that, in the control group, there were significantly less infants classified as normal in the third AIMS assessment than in the first (Z=-2.71, p=0.01).

**Table 3.** Classification of the preterm infants in the three groups according to the Alberta Infant Motor Scale

| Assessment | AIMS classification | Total<br>group<br>(n = 30)<br>Mean±SD | Control<br>group<br>(n = 11)<br>Mean±SD | Prone<br>group<br>(n = 9)<br>Mean±SD | Supine<br>group<br>(n = 10)<br>Mean±SD | FET | p |
| --- | --- | --- | --- | --- | --- | --- | --- |
| First | Normal | 30 | 11 | 9 | 10 |  |  |
|  | Suspicious | 0 | 0 | 0 | 0 | a) | a) |
|  | Abnormal | 0 | 0 | 0 | 0 |  |  |
| Second | Normal | 21 | 8 | 6 | 7 |  |  |
|  | Suspicious | 8 | 2 | 3 | 3 | 2.32 | 0.96 |
|  | Abnormal | 1 | 1 | 0 | 0 |  |  |
| Third | Normal | 16 | 3 | 5 | 8 |  |  |
|  | Suspicious | 9 | 7 | 2 | 0 | 10.46 | 0.02 |
|  | Abnormal | 5 | 1 | 2 | 2 |  |  |
| Fourth | Normal | 17 | 5 | 5 | 7 |  |  |
|  | Suspicious | 5 | 3 | 1 | 1 | 20.06 | 0.79 |
|  | Abnormal | 8 | 3 | 3 | 2 |  |  |

Note. FET: Fischer's Exact Test; a) No statistics were computed because AIMS classification is constant.

Figure 1 the trajectories of development of the different infants, displaying the changes of the final score in Alberta scale, in absolute values, along the four evaluations. The variability of the AIMS scores increases as infants grow older. In the first two evaluations, that all the preterm infants showed a motor development closer to the normal average than in the third and fourth evaluations, where AIMS values became much more disperse.

**Fig. 1.**
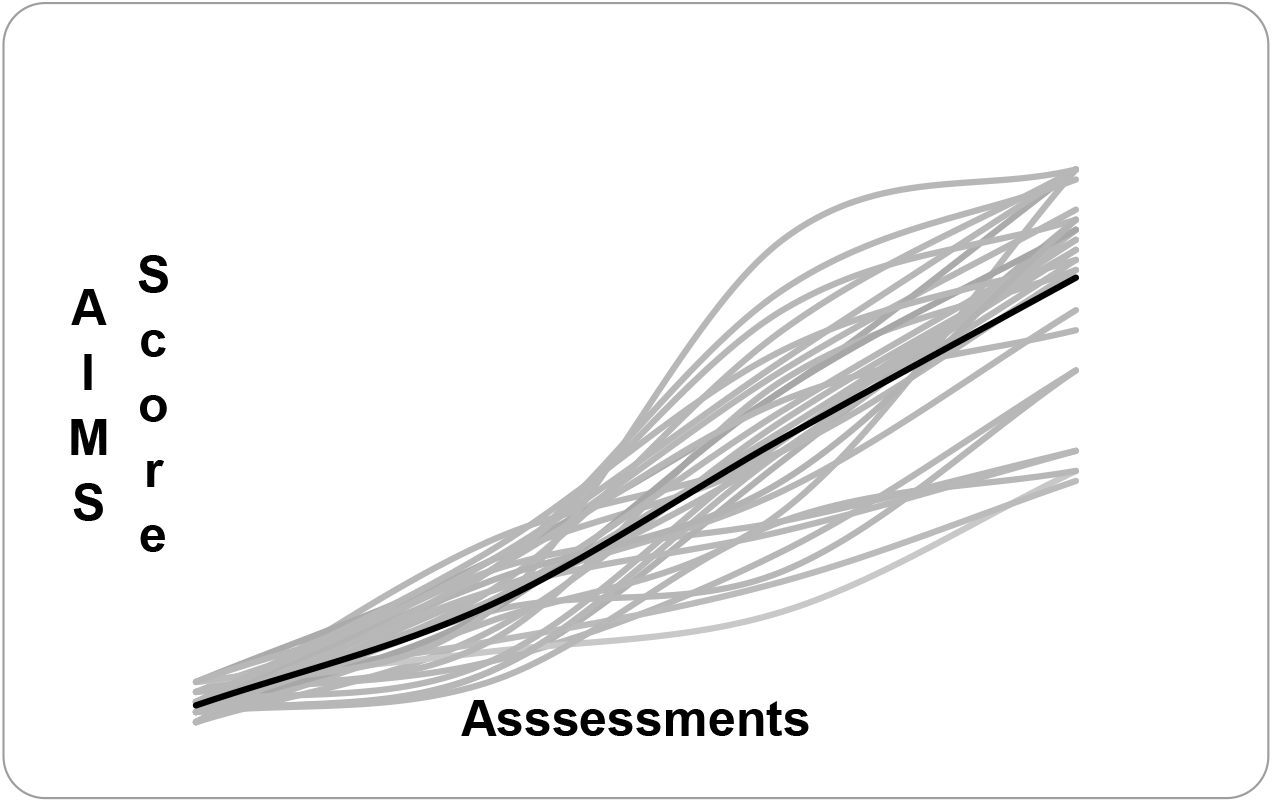
Final Alberta Infant Motor Scale (AIMS) score of the 30 preterm infants in the four assessments. Friedman test. Wilcoxon post-hoc test.

The results of the stepwise multiple linear regressions identified different predictors of performance in the AIMS, depending on the moment of the motor assessment. Variables entered as possible predictors were: maternal age, gestational age, birth weight, birth length, head circumference, chest circumference, APGAR (first and fifth minutes), days with mechanical ventilation, days with breathing support (O2), days with continuous positive airway pressure, length of hospital stay, weight when the infant left the ICU, and weight when the infant left the hospital. No predictors were found for the first assessment. For the other three assessments the following predictors were found: second assessment - length of hospital stay (Beta=-0.52, p=0.02; R^2^=0.27); third assessment - birth weight (Beta=0.51, p=0.02; R^2^=0.26); forth assessment - maternal age (Beta=0.54, p=0.01; R^2^=0.29).

The analysis of the phone call records made during follow-up of the mothers showed that only two mothers (22.2%) of the prone group reported to have followed the instructions correctly at all times. The remaining mothers reported not to have followed the instructions because of fear of leaving the child in prone, because the child did not stay in the prone position or cried, because of lack of time since they had to care for the other children, and because they forgot to follow the instructions. Thus, most mothers did not follow the instructions on prone positioning. On the other hand, most of the mothers of the supine group (70%) correctly followed the instructions.

## Discussion

The results showed that preterm infants achieved the motor developmental milestones within the first year of life. However, a larger number of infants were classified as suspicious or abnormal by the AIMS between 8 months and one year of corrected age. The strategy of providing guidance for caregivers was used so that preterm infants would receive motor stimulation at home. The number of infants with abnormal and suspicious motor development was higher in the group whose caregiver did not receive any guidance on motor stimulation compared to infants whose mothers received instructions.

Maternal guidance and training on the stimulation of motor development in preterm infants have been shown to result in positive outcomes for children. According to these authors, the results suggest that the support given to mothers may have contributed to their self-confidence, improving their ability to care for their children, and that the effective participation of mothers based on the instructions offered may have influenced the affective relationship with the infant, providing a greater stimulus for neurosensorimotor acquisition of the infant.^18^

In the present study, the 30 infants were classified as extremely preterm and very low birth weight, a fact explaining the mean length of hospital stay of 43.9 days for special care. In this respect, prolonged hospitalization is known to limit closer involvement of the parents with their infants. Another study involving very low birth weight preterm infants reported a mean length of stay of 60 days. According to the authors, the need for hospitalization of the infant alters family dynamics in an unexpected way since the parents have to deal with the emotions of hospitalization of the debilitated child. Possible complications may arise with the organization of daily routines to accompany the infant during hospitalization and with the preparation for discharge when the care will become the full responsibility of the parents.^19^

In an attempt to identify strategies designed to improve the development of preterm infants during longitudinal and home follow-up, the stimulation of body positioning has gained the attention of professionals as a protective practice for children. Prone positioning has shown some advantages for the cardiorespiratory system ^20^ and for the child’s motor development. A recent study demonstrated that children who cannot maintain the prone position on extended arms have a disadvantage, exhibiting delayed development.^21^

Placing the infant in the prone position during waking hours is an uncommon practice among Brazilian mothers.^8,22^ However, further studies are necessary to understand the reason for this parental practice. The present findings agree with previous studies reporting that many patients avoid placing their children in the prone position for play because the child is intolerant to this position, as demonstrated by crying. Consequently, the child is rarely exposed to this position.^10,23^ These findings corroborate another study showing that 44% of the infants were intolerant to the prone position and spent less than 15 minutes in this position for play during their waking hours.^24^

We understand that the time in the prone position is an important part of the daily routine of an infant and can be performed at different times over the day. Professionals must explain caregivers that the child may not tolerate the prone position on the first occasion and that the time can be gradually increased with increasing tolerance. To encourage the use of this position, the parents may talk and sing to the infant, use toys and mirrors, and maintain visual contact,^25^ reinforcing a playful way as instructed in this study.

Although the mothers of this study did not follow the prescription correctly, the importance of guidance on body positioning for the child’s motor development should be reinforced in public child care programs so that it can be better understood and applied by caregivers. The present results showed no significant difference between all associations of the positions. However, from a clinical point of view, we emphasize that 63.1% of the children whose mothers received guidance on body positioning achieved normal motor development according to the AIMS in the first year of life. The percentage was only 45.4% in the group that did not receive any guidance, with a difference of 17.7% between the intervention and control groups.

At 12 months of chronological age, an association was observed between AIMS scores and the environmental variable in the group of preterm infants studied. The older the mother, the better was the motor development of the infant. Our findings agree with a study showing that older parents have greater knowledge of infant development, favoring the development of their child.^26^ Another study following up 561 infants until 18 months of age demonstrated an association of environmental factors with motor development, especially in the second half of the first year of life.^27^ The evidence reinforces the need for professional guidance on child development. On the other hand, the association of birth weight and length of hospital stay with AIMS scores was no longer observed at 12 months corrected age of the infants.

### Limitations

This study has some limitations. The prescription of body positioning was not completely followed by the caregiver, a fact that can negatively influence the outcome. However, empowering caregivers is extremely important for the achievement of motor skills. In addition, the number of infants was lower than that estimated at the beginning of the study. The main reason was the difficulty in recruiting infants that met the inclusion criteria over the period studied. Obtaining a larger number of infants than those reported in this study is not simple since the study involved preterm newborns with exclusion factors and one-year follow-up by the same professional.

### Implications for Occupational Therapy Practice

The literature suggests an effect of body positioning on motor development. Evidence of the influence of body positioning after discharge of preterm infants is limited.

- Considering the increasing survival of preterm infants, guidance on body positioning could be a protective strategy for motor development to help mothers care.
- By the end of the first year of life most infants in the experimental groups (supine and prone positions) achieved a normal motor development, which did not happen in the control group (without position orientation).
- This study findings show evidences that mother orientation about child body position can help in preterm infants motor development.

### Conclusion

The present results show that prone positioning of preterm infants is little used at home and suggest that achieving motor developmental milestones involves body positioning in the first year of life. Home strategies for the care of this vulnerable population must be designed to achieve the best development as a protective factor.

## Data Availability

All data produced in the present work are contained in the manuscript.

## Acknowledgments

We thank the employees and families of the High-Risk Children Outpatient Clinic for their collaboration with this study.

## References

1. Glass HC, Costarino AT, Stayer SA, et al. Outcomes for extremely premature infants. Anesthesia and analgesia, 2015; 120(6), 1337–1351. https://doi:10.1213/ANE.0000000000000705

2. Blencowe, H., Cousens, S., Chou, D., Oestergaard, M., Say, L., Moller, A. B., Kinney, M., & Lawn, J. (2013). Born too soon: the global epidemiology of 15 million preterm births. Reproductive health, 10(1), S2. https://doi.org/10.1186/1742-4755-10-S1-S2

3. Blencowe, H., Lee, A. C., Cousens, S., Bahalim, A., Narwal, R., Zhong, N., Chou, D, Say, L, Modi, N, Katz, J, Vos, T, Marlow, N, &., Lawn, J. E. (2013). Preterm birth–associated neurodevelopmental impairment estimates at regional and global levels for 2010. Pediatric research, 74(S1), 17–34. https://doi:10.1038/pr.2013.204

4. Behrman, R. E. & Butler, A. S.(Eds.). (2007). Preterm birth: causes, consequences, and prevention. National Academies Press. Retrieved from https://www.ncbi.nlm.nih.gov/books/NBK11362/.

5. Liu, L., Oza, S., Hogan, D., Perin, J., Rudan, I., Lawn, J. E., Cousens S2, Mathers C & Black, R. E. (2015). Global, regional, and national causes of child mortality in 2000–13, with projections to inform post-2015 priorities: an updated systematic analysis. The Lancet, 385(9966), 430–440. https://doi.org/10.1016/S0140-6736(14)61698-6

6. World Health Organization. (2012). Born too soon: the global action report on preterm birth. Retrieved from http://www.who.int/pmnch/media/news/2012/201204_borntoosoon-report.pdf.

7. Darrah, J., Hodge, M., Magill-Evans, J., & Kembhavi, G. (2003). Stability of serial assessments of motor and communication abilities in typically developing infants— implications for screening. Early human development, 72(2), 97–110. https://doi.org/10.1016/S0378-3782(03)00027-6

8. Gontijo, A. P. B., de Castro Magalhães, L., & Guerra, M. Q. F. (2014). Assessing gross motor development of Brazilian infants. Pediatric Physical Therapy, 26(1), 48–55. https://doi:10.1097/PEP.0000000000000014

9. Salls, J. S., Silverman, L. N., & Gatty, C. M. (2002). The relationship of infant sleep and play positioning to motor milestone achievement. American Journal of Occupational Therapy, 56(5), 577–580. https://doi:10.5014/ajot.56.5.577

10. Dudek-Shriber, L., & Zelazny, S. (2007). The effects of prone positioning on the quality and acquisition of developmental milestones in four-month-old infants. Pediatric Physical Therapy, 19(1), 48–55. https://doi:10.1097/01.pep.0000234963.72945.b1

11. Santos, D. C., Gabbard, C., & Goncalves, V. M. (2000). Motor development during the first 6 months: the case of Brazilian infants. Infant and Child Development: An International Journal of Research and Practice, 9(3), 161–166. https://doi.org/10.1002/1522-7219(200009)9:3<161::AID-ICD229>3.0.CO;2-7

12. Lopes, V. B., de Lima, C. D., & Tudella, E. (2009). Motor acquisition rate in Brazilian infants. Infant and Child Development: An International Journal of Research and Practice, 18(2), 122–132. https://doi.org/10.1002/icd.595

13. Pinto, P. A. F., Falci, D. M., & Morais, R. (2017). Percepção, conhecimento e prática de pediatras quanto ao posicionamento do lactente e o desenvolvimento motor. Revista Pesquisa em Fisioterapia, 7(2), 149–156. https://doi.org/10.17267/2238-2704rpf.v7i2.1266

14. Sociedade Brasileira de Pediatria. (2012). Departamento Científico de Neonatologia, editor. Seguimento Ambulatorial do Prematuro de Risco.

15. Rugolo, L. M. S. D. S. (2005). Growth and developmental outcomes of the extremely preterm infant. Jornal de pediatria, 81(1), S101–S110. https://doi.org/10.1590/S0021-75572005000200013

16. Van Haastert, I. C., De Vries, L. S., Helders, P. J. M., & Jongmans, M. J. (2006). Early gross motor development of preterm infants according to the Alberta Infant Motor Scale. The Journal of pediatrics, 149(5), 617–622. http://doi.org/10.1016/j.jpeds.2006.07.025

17. Piper, M. C., Pinnell, L. E., Darrah, J., Maguire, T., & Byrne, P. J. (1992). Construction and validation of the Alberta Infant Motor Scale (AIMS). Canadian journal of public health= Revue canadienne de sante publique, 83, S46–50

18. Kayenne Formiga, C., Silva Pedrazzani, E., Pereira dos Santos Silva, F., & de Lima, C. D. (2004). Effectiveness of the early intervention program with preterm infants. Paidéia, 14(29), 301–311. http://dx.doi.org/10.1590/S0103-863x2004000300006

19. Sassá, A. H., & Silva Marcon, S. (2013). Evaluation of families of infants with very low birth weight in home care. Texto & Contexto Enfermagem, 22(2),442–451. http://dx.doi.org/10.1590/S0104-07072013000200021

20. Oishi, Y., Ohta, H., Hirose, T., Nakaya, S., Tsuchiya, K., Nakagawa, M., Kusakawa, I, Sato, T, Obonai, T, Nishida, H & Yoda, H. (2018). Combined effects of body position and sleep status on the cardiorespiratory stability of near-term infants. Scientific reports, 8(1), 8845. https://doi:10.1038/s41598-018-27212-8

21. Senju, A., Shimono, M., Tsuji, M., Suga, R., Shibata, E., Fujino, Y., Kawamoto, T, & Kusuhara, K. (2018). Inability of infants to push up in the prone position and subsequent development. Pediatrics International. http://doi.org/10.1111/ped.13632

22. Silva, P. L., Santos, D. C. C., & Gonçalves, V. M. G. (2006). Influence of child-rearing practices on infant’s motor development between the sixth and twelfth months of life. Brazilian Journal of Physical Therapy, 10(2),225–231.

23. Valentini, N. C., & Saccani, R. (2012). Brazilian Validation of the Alberta Infant Motor Scale. Physical therapy, 92(3), 440–447. http://doi.org/10.2522/ptj.20110036

24. Zachry, A. H., & Kitzmann, K. M. (2011). Caregiver awareness of prone play recommendations. American Journal of Occupational Therapy, 65(1), 101–105. http://doi:10.5014/ajot.2011.09100

25. Coulter-O’Berry, C., & Lima, D. (2006). Tummy time tools: Activities to help you position, carry, hold and play with your baby. Retrieved from <https://www.choa.org/~/media/files/Childrens/medical-services/orthopaedics/orthotics-and-prosthetics/tummy-time-tools-update-2014.pdf?la=en>.

26. Lima, L. N., Vale-Dias, M. D. L., & Mendes, T. F. V. (2012). Crenças parentais sobre o desenvolvimento da criança e sua relação com o cuidar. Revista INFAD, 1(1),53–62.

27. Saccani, R., Valentini, N. C., Pereira, K. R., Müller, A. B., & Gabbard, C. (2013). Associations of biological factors and affordances in the home with infant motor development. Pediatrics International, 55(2), 197–203. https://doi.org/10.1111/ped.12042

